# The Promise and Peril of Large Language Models in Digital Health: GPT-4 Personalizes Cardiovascular Patient Education but Amplifies Gender Biases

**DOI:** 10.1101/2025.11.19.25340616

**Authors:** Samah Khan, Georgeta Vaidean

## Abstract

**Background:** Gender-neutral patient education materials often overlook critical sex-based differences in cardiovascular disease (CVD). Large Language Models (LLMs) like GPT-4 offer a potential tool for personalizing health communication, but their ability to correct gender gaps without introducing new biases is unknown.

**Methods:** We identified seven publicly available English-language CVD prevention handouts from major health organizations. Using GPT-4 API in August 2025, we generated gender-specific revisions for a 55-year-old male and female audience via standardized prompts. We provided structured prompts instructing the model to include evidence-based, sex-specific risk factors and symptoms. Original and revised materials were evaluated using Flesch-Kincaid Reading Ease, a novel 10-point gender-inclusivity checklist, and qualitative thematic analysis.

**Findings:** GPT-4 revisions substantially improved gender-inclusivity scores (Original median: 3.0/10, IQR=0.0-3.0; Male-tailored median: 8.0, IQR=7.0-9.0; Female-tailored median: 10.0, IQR=10.0-10.0). Readability was maintained. However, qualitative analysis revealed that while female-tailored versions excelled at incorporating biological facts (e.g., menopause), male-tailored versions often missed key clinical factors. For instance, 4 of 7 revisions failed to mention erectile dysfunction as a CVD risk marker. Revisions also occasionally relied on social stereotypes (e.g., “bottling up emotions”). Furthermore, both versions showed evidence of linguistic bias (framing female symptoms as ‘atypical,’ thereby reinforcing the male-centric clinical paradigm) and gendered assumptions in recommended activities.

**Conclusion:** LLMs can rapidly improve gender-specificity in patient education but can also perpetuate harmful stereotypes and linguistic biases likely absorbed from their training data. Their use requires careful, critical oversight to avoid undermining and to potentially advance health equity.

**Author Summary:** *Why was this study done?:* Heart disease affects men and women differently, but most patient education materials tend to be the same for everyone. We wanted to see if Artificial Intelligence, specifically large language models like GPT-4, could help rewrite these materials to be more accurate and helpful for each gender.

*What did the researchers do and find?:* We took seven heart disease prevention online handouts from major public health and clinical organizations. We used GPT-4 to create new versions specifically for men and for women, including male-specific risk markers (e.g., erectile dysfunction) and female-specific risk enhancers (e.g., menopause). We found that the original handouts were not very gender-specific. The AI-revised versions were dramatically better, especially for women, providing more relevant information without making the text harder to read. However, we also found the AI sometimes made mistakes, like using stereotypes about how men handle emotions.

*What do these findings mean?:* This means AI can be a powerful assistant for creating drafts, but it’s not a replacement for human expertise. To be used safely, a healthcare professional must always check the AI’s work to ensure it is medically accurate and free from harmful stereotypes.

## Introduction

Cardiovascular disease (CVD) remains the foremost cause of mortality globally, yet its manifestation, risk profiles, and optimal prevention strategies exhibit significant variation between sexes [1, 2]. For instance, women are more likely to present with “atypical” symptoms such as fatigue, nausea, or back pain during a myocardial infarction, often leading to delays in diagnosis and treatment [3, 4]. Biological factors, including the cardioprotective role of endogenous estrogen pre-menopause and its decline thereafter, fundamentally alter women’s CVD risk trajectory [5]. Conversely, men often develop CVD at a younger age and face distinct risk markers, such as erectile dysfunction [6].

Despite these well-documented differences, a substantial portion of patient-facing educational materials on CVD prevention remains gender-neutral [7]. This “one-size-fits-all” approach, while simplifying content creation, risks flattening critical nuances, potentially leaving patients ill-informed about their specific risks and symptoms. The field of health literacy has long advocated for tailored communication to improve comprehension and self-efficacy [8], yet creating multiple versions of materials is a resource-intensive process.

The emergence of Large Language Models (LLMs) like GPT-4 offers a potential paradigm shift. These models can generate human-like text and follow complex instructions, presenting an unprecedented opportunity to automate the personalization of health information at scale [9]. Preliminary research has explored LLMs for improving readability and simplifying medical jargon [10], but their ability to address nuanced issues of gender representation and equity in clinical content remains largely unexplored.

We hypothesized that GPT-4 can be strategically prompted to revise existing CVD prevention materials into gender-specific versions that are both highly readable and significantly more inclusive of evidence-based sex differences. Specifically, we aimed to:

1. Quantify the extent of gender-specific content in widely used patient education handouts, and
2. Evaluate the efficacy of GPT-4 in enhancing gender representation while preserving or improving readability.

Our study therefore aims to serve as a critical, real-world evaluation of a prominent AI tool for a core digital health application: the scalable personalization of patient-facing communication, with further potential to support more equitable health education materials.

## Results

### 1. Quantitative Improvements in Gender-Specificity and Readability

The seven original patient education handouts demonstrated moderate readability (Median Flesch Reading Ease = 64.0, IQR = 60.7-71.2). However, their performance on the gender-inclusivity checklist was poor (Median Score = 3.0, IQR = 0.0-3.0). Notably, materials from the CDC and NHS scored 0/10, containing no sex-specific content. MedLine Plus scored highest (6/10) but still omitted key differentiators like symptom variation. No original material explicitly discussed hormonal influences like testosterone’s role in male cardiovascular health or pregnancy-related risks for women.

GPT-4 revision led to a dramatic and consistent improvement in gender-inclusivity scores for both male- and female-tailored versions (Figure 1). The increase was most pronounced for female-tailored materials, which achieved a perfect median score of 10.0 (IQR = 10.0-10.0). All seven female-tailored versions addressed female-predominant symptoms, menopause-related risk, and hormonal influences. Male-tailored versions also showed significant improvement (Median Score = 8.0, IQR = 7.0-9.0), though they occasionally omitted specific factors like erectile dysfunction.

**Table 1.** Readability and Gender-Inclusivity Scores for Original and GPT-4 Revised Patient Education Materials.

| Source | Flesch Reading Ease |  |  | Gender-Inclusivity Score (0-10) |  |  |
| --- | --- | --- | --- | --- | --- | --- |
|  | Original | Male-Tailored | Female-Tailored | Original | Male-Tailored | Female-Tailored |
| Mayo Clinic | 71.4 | 73.2 | 73.6 | 3 | 9 | 10 |
| AHA | 58.5 | 68.8 | 70.3 | 2 | 7 | 10 |
| Cleveland Clinic | 68.7 | 73.5 | 67.8 | 3 | 8 | 10 |
| CDC | 63 | 62.5 | 65.1 | 0 | 9 | 10 |
| NHS | 60.7 | 71.4 | 75.9 | 0 | 7 | 9 |
| UC Davis | 64 | 78.8 | 68.1 | 3 | 7 | 10 |
| MedLine Plus | 71.2 | 75.6 | 73.8 | 6 | 9 | 10 |
| Median (IQR) | 64.0 (60.7-71.2) | 73.2 (68.8-75.6) | 70.3 (67.8-73.8) | 3.0 (0.0-3.0) | 8.0 (7.0-9.0) | 10.0 (10.0-10.0) |

**Figure 1.**
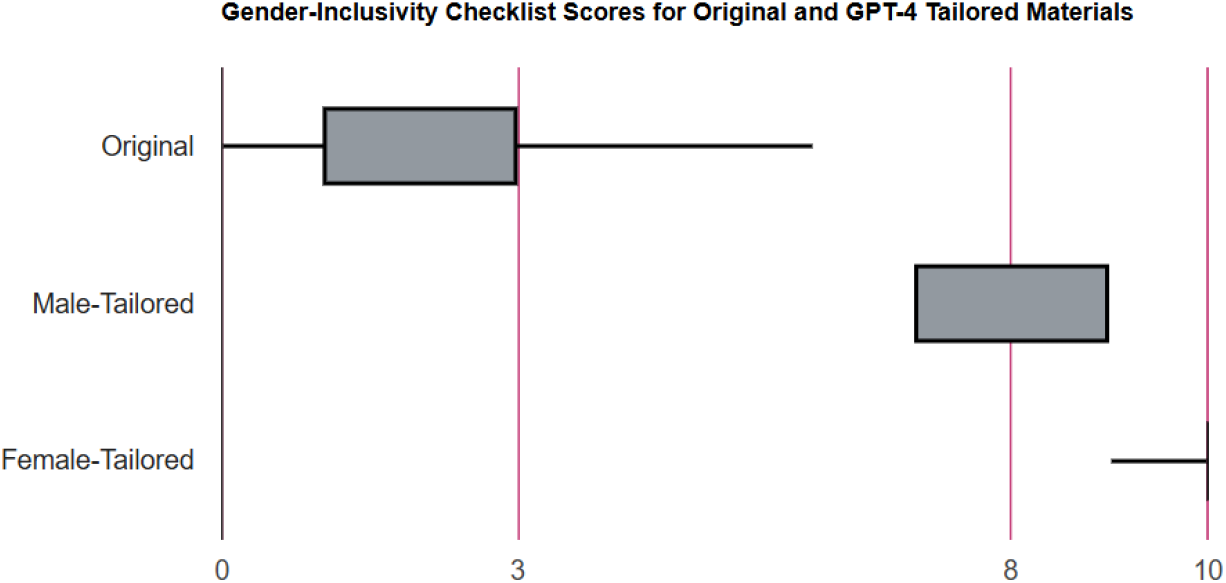
**Boxplot of gender-inclusivity checklist scores (0-10 scale) for original, male-tailored, and female-tailored patient education materials. The female-tailored condition demonstrates near-perfect performance, with a median score of 10 and a tight interquartile range (IQR) at the maximum value. A single score of 9 falls below the IQR, indicated by the whisker**.

A Friedman test revealed a statistically significant difference in gender-inclusivity scores across the three conditions (χ^2^(2) = 14.0, p < 0.001). Post-hoc Nemenyi tests confirmed that both male-tailored (p = 0.020) and female-tailored (p < 0.001) scores were significantly higher than original scores. The difference between male and female-tailored scores was not statistically significant (p = 0.47).

GPT-4 revisions not only enhanced gender-specificity but also maintained high readability. The median Flesch Reading Ease score was 73.2 (IQR = 68.8-75.6) for male-tailored and 70.3 (IQR = 67.8-73.8) for female-tailored versions. A separate Friedman test found no statistically significant difference in readability scores across the three conditions (χ^2^(2) = 5.43, p = 0.066), indicating that the process of adding complex, specific information did not compromise textual accessibility.

### 2. Qualitative Analysis Reveals Persistent Stereotypes and Linguistic Biases

A qualitative analysis of the GPT-4 outputs uncovered instances where the model amplified societal biases.

- **Linguistic Framing of Male-as-Default**: In describing symptoms, the NHS female revision noted women experience “‘less common’ heart attack symptoms.” This framing positions the male clinical presentation as the norm and the female presentation as a deviation, potentially reinforcing diagnostic disparities.
- **Omission of Male-Specific Biology**: Despite explicit prompting, most male-tailored revisions failed to mention erectile dysfunction as a CVD risk marker or discuss the role of testosterone.
- **Reinforcement of Social Stereotypes**: The model occasionally substituted clinical facts with social tropes. For example, a male revision generated by GPT-4 stated, “Men are more likely to bottle up emotions, which can lead to stress-related health problems,” a non-evidence-based generalization.
- **Gendered Assumptions in Preventive Actions**: Recommendations for physical activity and overcoming barriers often reflected traditional gender roles. For example, when a gender-neutral original handout listed ‘housekeeping’ as an example of physical activity, this was retained only in its female-tailored GPT-4 revision. In contrast, a male-tailored revision of a different source suggested ‘affordable gyms,’ demonstrating how the model imposed gendered assumptions on neutral content.

## Discussion

### 4.1. Principal Findings

Our study suggests that GPT-4 was highly effective at integrating evidence-based, sex-specific information into CVD prevention materials, with a particularly strong performance in creating content for women. This likely reflects a more substantial corpus of literature and public health messaging addressing the historical neglect of women’s cardiovascular health. The improvement in male-tailored content, while significant, was less complete, suggesting that nuanced men’s health topics are less prominently featured in the model’s training data.

### 4.2. Implications of Bias and Stereotype Perpetuation

Our most critical finding is that the quantitative success of LLMs in adding content can mask profound qualitative failures in equity. GPT-4 did not merely reflect biases present in its training data; in several instances, it actively generated and amplified them. The consistent omission of erectile dysfunction as a CVD risk marker, despite explicit male-context prompting, constitutes a critical clinical oversight and reveals a gap in the model’s assimilation of men’s health literature. Phrases like “bottling up emotions” replace biomedical fact with social stereotypes. The description of female symptoms as “less common” is not merely a semantic issue but a manifestation of the androcentric bias historically prevalent in medicine, which can have tangible consequences for women’s care [11].

These biases likely stem from the model’s training on vast datasets that contain societal and medical biases. Without explicit correction, the model replicates these patterns. This indicates that using LLMs for health equity purposes should not be a passive process. It requires active, critical prompting and oversight.

### 4.3. Limitations and Future Directions

This study has several limitations. The sample size of seven handouts, while diverse, is small. The evaluation relied on an ad hoc checklist developed for this study, informed by established literature. Our evaluation, while incorporating a structured checklist and qualitative analysis, did not include patient perspectives on the perceived empathy, trustworthiness, or usefulness of the AI-generated materials. We did not test the materials with actual patients to assess comprehension or behavior change, a vital next step. Future research should explore LLM customization for non-binary individuals, adaptation into other languages, and application to other disease areas.

A key direction for future work is to develop and test “bias-aware” prompting strategies that explicitly instruct the model to avoid stereotypes and use patient-centric language. Furthermore, evaluation should include feedback from diverse patient groups to assess not just accuracy but also perceived empathy and relevance.

### 4.4. Conclusion

In conclusion, LLMs like GPT-4 represent a powerful but double-edged sword for creating equitable health education. They can rapidly generate more personalized content but can also codify existing biases. Therefore, their implementation must be guided by a framework of critical digital health literacy, where AI assists in drafting, but human expertise (particularly in ethics and health equity) remains essential for editing, validation, and ensuring the promotion of authentic patient-centered care.

## Materials and Methods

### 1. Material Selection

To simulate a patient’s information-seeking behavior, we conducted a systematic search using the Google search engine on August 17, 2025. The search query ‘how to prevent a heart attack’ was used. We included the top 7 unique English-language patient education handouts from the first page of results, representing major public health and clinical organizations (e.g., CDC, NHS, Mayo Clinic). This approach was chosen to capture the materials a typical patient is most likely to encounter during a common information-seeking behavior, thereby enhancing the external validity of our study. All original handouts are available in the OSF repository.

### 2. LLM Revision Process

Each handout was processed using the OpenAI GPT-4 API on August 17, 2025 with the specified prompts. The chat completion API was used in a temporary (stateless) session to ensure that each prompt was processed independently without influence from previous interactions. The phrase ‘where clinically established’ was included in the prompts to constrain the model’s output to only those sex-specific differences supported by robust clinical evidence, thereby minimizing the generation of unverified or speculative content. We used the GPT-4 model with a temperature setting of 0 to maximize determinism and reproducibility. Prompts were refined through an iterative piloting process with three handouts to establish reliability. These pilot handouts were not included in the final analysis of the seven primary handouts. All outputs were saved verbatim.

“Revise this heart disease prevention material for a 55-year-old male audience at a 6th-grade reading level. Where clinically established, explicitly include:

- Male-predominant symptom presentations
- Biological risk factors more common in men
- Prevention strategies with different efficacy/considerations for men
- Maintain strict medical accuracy and cite only evidence-based differences.”

“Revise this heart disease prevention material for a 55-year-old female audience at a 6th-grade reading level. Where clinically established, explicitly include:

- Female-predominant symptom presentations
- Biological risk factors more common in women
- Prevention strategies with different efficacy/considerations for women
- Maintain strict medical accuracy and cite only evidence-based differences.”

### 3. Evaluation Metrics

A novel 10-point gender-inclusivity checklist was developed de novo based on a comprehensive review of literature on sex-based differences in CVD [12, 13, 14, 15, 16, 17]. It consisted of 5 domains: (1) sex-specific symptoms, (2) biological risk factors, (3) hormonal influences, (4) sex-specific risk markers (e.g., erectile dysfunction), and (5) considerations for prevention strategies. Each domain was scored 0 (absent), 1 (mentioned but incomplete), or 2 (clearly and accurately described), for a total of 10 points. All materials were scored by a single reviewer using this structured, criteria-based checklist to ensure consistency in application. The objective nature of the checklist items (e.g., presence/absence of a specific risk factor) was designed to minimize subjective interpretation.

### 4. Data Analysis

Descriptive statistics (medians and interquartile ranges (IQR)) were used to summarize readability and gender-inclusivity scores. Given the small sample size (N=7 handouts) and the ordinal nature of the checklist scores, the non-parametric Friedman test was used to compare gender-inclusivity scores across the three revision types (Original, Male-tailored, Female-tailored). A Friedman test was also performed on the Flesch Reading Ease scores. A statistically significant Friedman test was followed by post-hoc pairwise comparisons. Statistical analysis was performed using StatsKingdom.com. A p-value of < 0.05 was considered statistically significant.

## Data Availability Statement

All data and materials are available in the OSF repository: https://doi.org/10.17605/OSF.IO/YNA6K

